# A large outbreak of COVID-19 in a UK prison, October 2020 to April 2021

**DOI:** 10.1101/2022.02.02.22269960

**Authors:** James P. Adamson, Christopher Smith, Nicole Pacchiarini, Thomas Richard Connor, Janet Wallsgrove, Ian Coles, Clare Frost, Angharad Edwards, Jaisi Sinha, Catherine Moore, Steph Perrett, Christie Craddock, Clare Sawyer, Alison Waldram, Alicia Barrasa, Daniel Rh. Thomas, Philip Daniels, Heather Lewis

## Abstract

**Introduction:** Prisons are susceptible to outbreaks. Control measures focusing on isolation and cohorting negatively affect wellbeing. We present an outbreak of COVID-19 in a large male prison in Wales, UK, October 2020 to April 2021, and discuss control measures.

**Methods:** We gathered case-information, including demographics, staff-residence postcode, resident cell number, work areas/dates, test results, staff interview dates/notes and resident prison-transfer dates. Epidemiological curves were mapped by prison location. Control measures included isolation (exclusion from work or cell-isolation), cohorting (new admissions and work-area groups), asymptomatic testing (case-finding), removal of communal dining and movement restrictions. Facemask use and enhanced hygiene were already in place. Whole genome sequencing (WGS) and interviews determined genetic relationship between cases plausibility of transmission.

**Results:** Of 453 cases, 53% (n=242) were staff, most aged 25-34 years (11.5% females, 27.15% males) and symptomatic (64%). Crude attack-rate was higher in staff (29%, 95%CI: 26-64%) than in residents (12%, 95%CI: 9-15%).

**Conclusions:** Whole genome sequencing can help differentiate multiple introductions from person-to-person transmission in prisons. It should be introduced alongside asymptomatic testing as soon as possible to control prison outbreaks. Timely epidemiological investigation, including data visualisation, allowed dynamic risk assessment and proportionate control measures, minimising reduction in resident welfare.

## Introduction

Prisons are crowded communal settings. During the COVID-19 pandemic, prisons have been highly susceptible to outbreaks, resulting in high levels of morbidity and mortality in residents and staff [1–3]. During the first wave, 7.6 confirmed COVID-19 cases were reported per 1000 prison residents in England and Wales compared with 4.9 in the general population [4]. In the second wave, this rose to 75 cases per 1000 population in prisons, compared to 46 cases per 1000 overall, by the end of December 2020 [5]. Control measures focusing on isolation and cohorting were initiated in prisons in Wales [6], reducing mixing and visits. These restrictive regimes can negatively affect physical and mental wellbeing through loss of control [7–9], consequently reducing cooperation: COVID-19 caused prison unrest and rioting in Europe early in the pandemic [10]. This is concerning given prisoners have worse physical and mental health than the general population and are regularly exposed to many health risks including smoking, poor hygiene and weakened immunity [3]. Measures to limit the spread and impact of COVID-19 in Welsh prisons began several weeks before the first prison-cases were seen. On 11 February 2020, Public Health England circulated their first interim guidance for COVID-19 in prisons, which was adopted in Wales. Infection control processes were established in all Welsh prisons, continuing throughout the pandemic in line with further guidance.

In October 2020, Public Health Wales (PHW) was notified of a case of COVID-19 in a resident of a large male prison in South Wales, UK (“Prison A”), which has approximately 1700 residents and 850 staff. This index case had a history of respiratory illness and was in hospital at the time of notification. After a negative PCR result on hospital admission, they tested PCR positive during their stay, classifying them as a hospital-acquired case. Two weeks later, an incident management team (IMT) was convened to review 25 more cases (20 residents, 5 staff), epidemiologically linked to the index case. The IMT declared an outbreak and established an outbreak control team (OCT) which managed this outbreak through collaborative decisions, as defined in the outbreak plan for Wales [11].

## Methods

The OCT met weekly to discuss case numbers, epidemiology, control measures and operational issues related to the outbreak. The roles and organisations of OCT members are given in Table 1. We describe the epidemiology and control measures implemented for this outbreak.

**Table 1.** Overview of roles of the OCT membership, 14 October 2020 - 21 April 2021

| Organization | Job Title | Role |
| --- | --- | --- |
| **Public Health Wales** | Consultant in Public Health | Chair |
|  | UK FETP Fellow | Epidemiological investigation |
|  | Improvements Manager | Line listing management & telephone interviews |
|  | Health Protection Nurse | Liaison with prison about cases |
|  | Lead Nurse for Health and Justice | Providing context at an all-Wales and UK level |
|  | Bioinformatician | WGS methods and results interpretation |
| **Local Health Board** | Consultant in Public Health | Providing local health board delivery information |
| **Prison A** | Director | Strategic & tactical intelligence about Prison A |
|  | Deputy Director | Strategic & tactical intelligence about Prison A |
|  | Head of Health Care | Intelligence on operational aspects of Prison A |
|  | Clinical Lead | Intelligence on operational aspects of Prison A |
|  | Compliance Manager | Detailed case information & operational aspects |
| **Local Council** | Environmental Health Officer | Local Government context |
| **HMPPS Wales** | Director (Strategic Support & Assurance) | Providing intelligence at a UK prisons level |

*UK FETP – UK Field epidemiology training programme*
*WGS – whole genome sequencing*
*HMPPS – Her Majesty's Prisons and Probation Service*

A possible case was any staff or resident at Prison A on or after 14 Oct 2020 who had symptoms compatible with COVID-19 (cough and/or fever and/or loss of smell/taste), without a PCR test result. A probable case was a possible case with a positive PCR test result that had not been verified by PHW. A confirmed case was any probable case with a positive PCR test result, which had been verified by PHW. A discarded case was a possible case whose PCR test result was negative.

In accordance to then-current national guidance, symptomatic residents and staff were tested for the presence of SARS-CoV-2 RNA using PCR tests. Asymptomatic screening was introduced in the Admissions unit (A-Block) from December 2020 (see control measures).

We telephoned all staff cases to discuss their movements and contacts prior to testing positive; no resident interviewing was done due to concerns of breaking self-isolation to access a telephone. Instead, intelligence on resident cases was obtained from prison staff. Given the restrictions of movement in place during this outbreak, this information was deemed accurate and reliable by the OCT.

Case data were managed using an Excel line list containing demographic information (age, sex), staff residence postcode, resident cell number, work areas/dates, laboratory results (test dates, result status, laboratory IDs, whole genome sequencing (WGS) links), interview dates/notes and resident prison-transfer dates. Line list management was performed by a single member of staff, in frequent contact with prison, “Test, Trace, Protect” (TTP – contact tracing) and laboratory colleagues.

### Epidemiological investigations

Cases were plotted on an epidemiological curve, then mapped by accommodation block and work location to show frequency and distribution over time (total and last 28 days). Prison A’s accommodation is divided into 10 accommodation blocks (box 1) and other functional areas including Industries, where the residents of working age undertake assigned work-activities, Laundry, Canteen, Gym and Healthcare.

Epidemiological curves and case location mapping were updated weekly, providing dynamic visual aids for risk assessment. Where cases were present in multiple locations, overlap was highlighted on the maps. Test date was used for asymptomatic cases and symptom onset date used for symptomatic cases. Crude attack rates were calculated by location using prison records for staff and residents (January 2021: 1693 staff and 832 staff). Variation in attack rates were measured using a two-proportion z-test. Finally, case hospitalisation ratios for staff and residents were calculated.

Whole genome sequencing was carried out by the PHW Pathogens Genomics Unit (PenGU) using the ARTIC protocol for “Illumina” (standard turnaround) or Oxford “Nanopore” (rapid turnaround) to assess the genetic relationship between cases, aid epidemiological investigation and determine control measures. Sequencing was performed only on samples meeting set quality criteria (including a diagnostic result with a cycle threshold value of less than 30). Samples were processed using the ncov2019 ARTIC Nextflow pipeline [12] and were analysed using the CLIMB COVID analysis platform [13] making use of the civet [14] and MircoReact [15] tools for analysis and visualisation of the outbreak.

## Results

Between October 2020 and April 2021 (189 days), Prison A reported 453 cases (staff n=242, 53%; residents n=211, 47%) (Table 2). Most were symptomatic (64%; staff n=163, 67%; residents n=126, 60%) (Table 3) with a mode of 25-34 years for both sexes (11.5% females, 27.15% males) (Figure 1). Three staff were hospitalised due to their COVID-19 illness; case hospitalisation ratios were 0.36 for staff and 0.06 for residents. One death was linked to this outbreak; a resident with a positive PCR test result more than 28 days but fewer than 90 days before death. Telephone interviews were completed with 99% of staff cases.

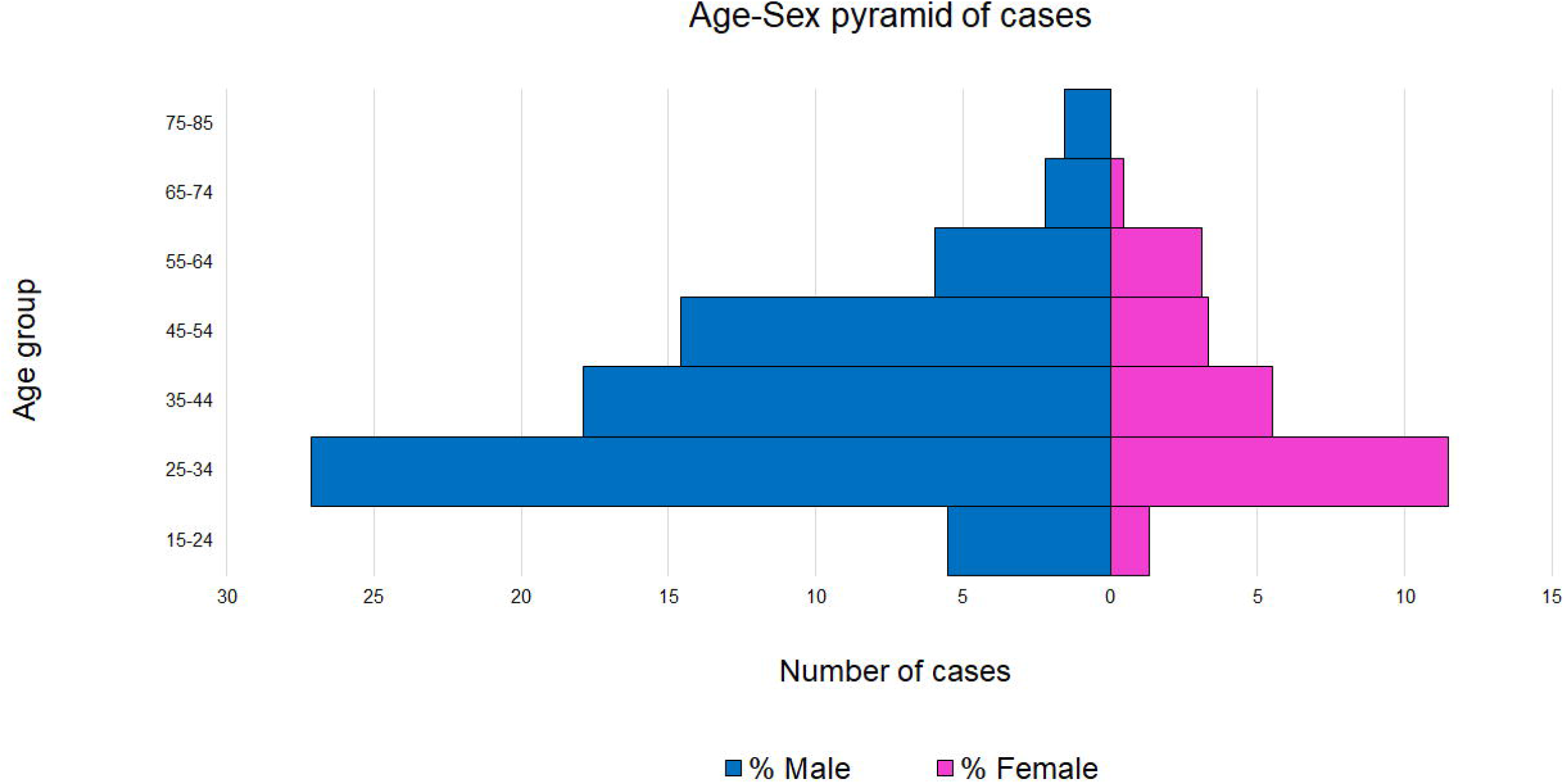

**Table 2.** Overview of cases by accommodation units, Prison A, January 2021 (N=332’)

| Area | Staff cases |  |  |  | Resident cases |  |  |  | Total cases |  |
| --- | --- | --- | --- | --- | --- | --- | --- | --- | --- | --- |
|  | Pop. | Symp. | Asymp. | AR* (%) | Pop. | Symp. | Asymp. | AR* (%) | n | AR* (%) |
| A | 48 | 22 | 9 | 64.6 | 355 | 24 | 65 | 25.1 | 120 | 29.8 |
| B | 42 | 10 | 3 | 31 | 366 | 9 | 0 | 2.5 | 22 | 5.4 |
| C | 17 | 3 | 1 | 23.5 | 74 | 0 | 0 | 0 | 4 | 4.4 |
| D | 14 | 8 | 0 | 57.1 | 91 | 23 | 1 | 26.4 | 32 | 30.5 |
| SCU | 10 | 8 | 1 | 90 | 12 | 0 | 1 | 8.3 | 10 | 45.5 |
| T | 78 | 15 | 10 | 32.1 | 410 | 36 | 4 | 9.8 | 65 | 13.3 |
| X | 47 | 10 | 4 | 29.8 | 325 | 14 | 11 | 7.7 | 39 | 10.5 |
| YPU | 44 | 17 | 21 | 86.4 | 60 | 0 | 2 | 3.3 | 40 | 38.5 |
| **Total** | **300** | **93** | **49** | **-** | **1693** | **106** | **84** | **-** | **332** | **-** |

*1 - Note: some staff worked in more than one location during this outbreak, so total is greater than 242*
*Pop – population; Symp. – symptomatic; Asympt. – asymptomatic; AR – attack rate*

**Table 3.**
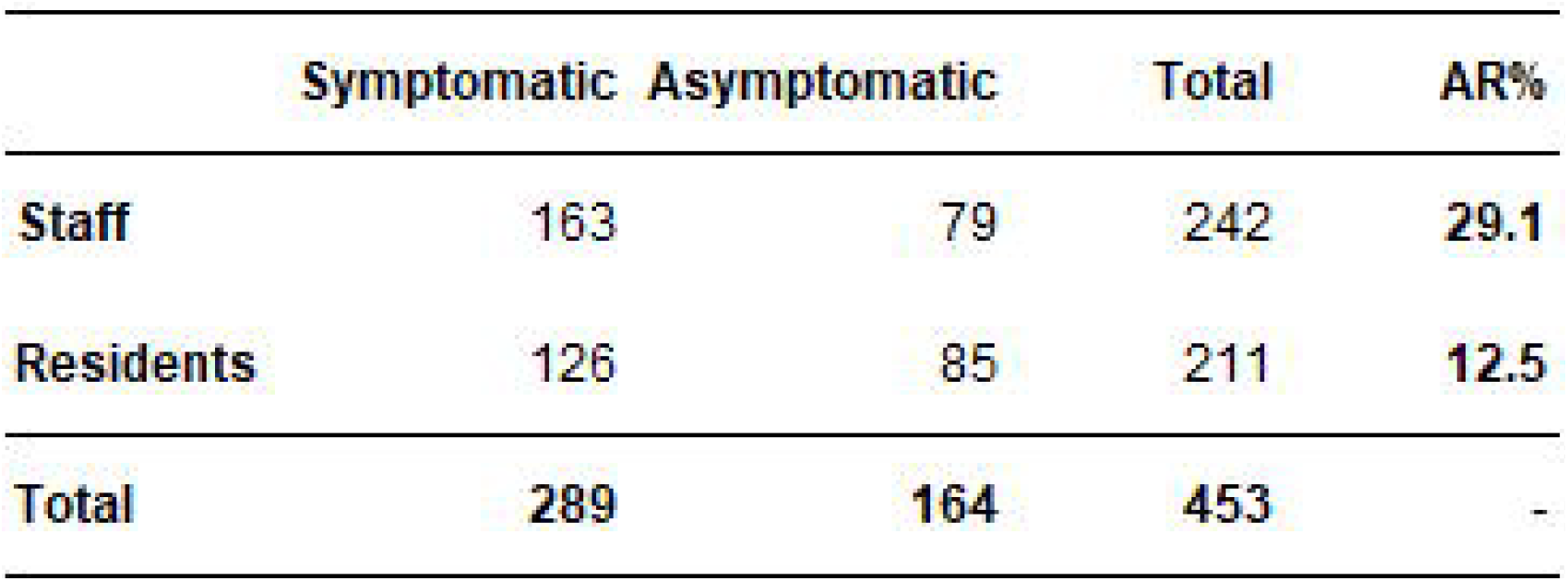
Overview of case types at Prison A, 14 October 2020 - 21 April 2021 (N=453)

|  | **Symptomatic** | **Asymptomatic** | **Total** | **AR%** |
| --- | --- | --- | --- | --- |
| **Staff** | 163 | 79 | 242 | **29.1** |
| **Residents** | 126 | 85 | 211 | **12.5** |
| **Total** | **289** | **164** | **453** | **-** |

The index case was resident in A-block prior to hospital admission; 12 staff cases worked hospital bed watch shifts for the index case during their infectious period and subsequently worked in A-block, T-block, young persons’ unit (YPU) and safer custody unit (SCU). However, there were already staff cases in SCU prior to these staff working back in Prison A.

Crude attack rate was higher in staff (29%, 95%CI: 26-64%) than in residents (12%, 95%CI: 9-15%). Accommodation-units’ specific attack rates ranged from 0% to 26% in residents and 24% to 90% in staff (table 2). Admissions (A-block), D-block, SCU and YPU experienced the highest overall attack rates (table 2). An overview of staff cases in other work locations (table 4) shows highest attack rates in the testing and mentor teams.

**Table 4.** Overview of staff cases by staff work location (L) or team (T), Prison A (N=103^†^), 14 October 2020 - 21 April 2021

| Location or Team | Cases | Pop.* | AR** % |
| --- | --- | --- | --- |
| Admin building and senior management team | 8 | 45 | 17.8 |
| Education and training teams | 7 | 71 | 9.9 |
| Gym | 4 | 15 | 26.7 |
| Healthcare | 8 | 49 | 16.3 |
| Interventions team | 2 | 25 | 8.0 |
| Mentor, families and pastoral support teams | 6 | 30 | 20.0 |
| Main industries | 9 | 20 | 45.0 |
| Main stores, facilities and waste management | 20 | 56 | 35.7 |
| Offender management unit | 7 | 59 | 11.9 |
| Security and nights teams | 18 | 61 | 29.5 |
| Testing team and pharmacy | 4 | 11 | 36.4 |
| Other*** | 10 | - | - |
| **Total** | **103** | **442** | **23.4** |

*1 - **Note** : some staff worked in more than one location during this outbreak.*
*(Some teams have been combined to maintain anonymity)*
*\* Pop. – population of staff location of team*
*\*\* AR – attack rate*
*\*\*\* Other contractors or short-term staff*

An epidemic curve (Figure 2), highlighting dates of key control measures, shows that the first half of the outbreak (87 days) comprised 346 (76%) of the cases reported. Of these, 75 (22%) were asymptomatic. During the second half of the outbreak (86 days), 82 of the 106 cases (77%) were asymptomatic. Mapping cases by area of work or residence showed variation in attack rates by area and which cases were associated to more than one location (Figure 3).

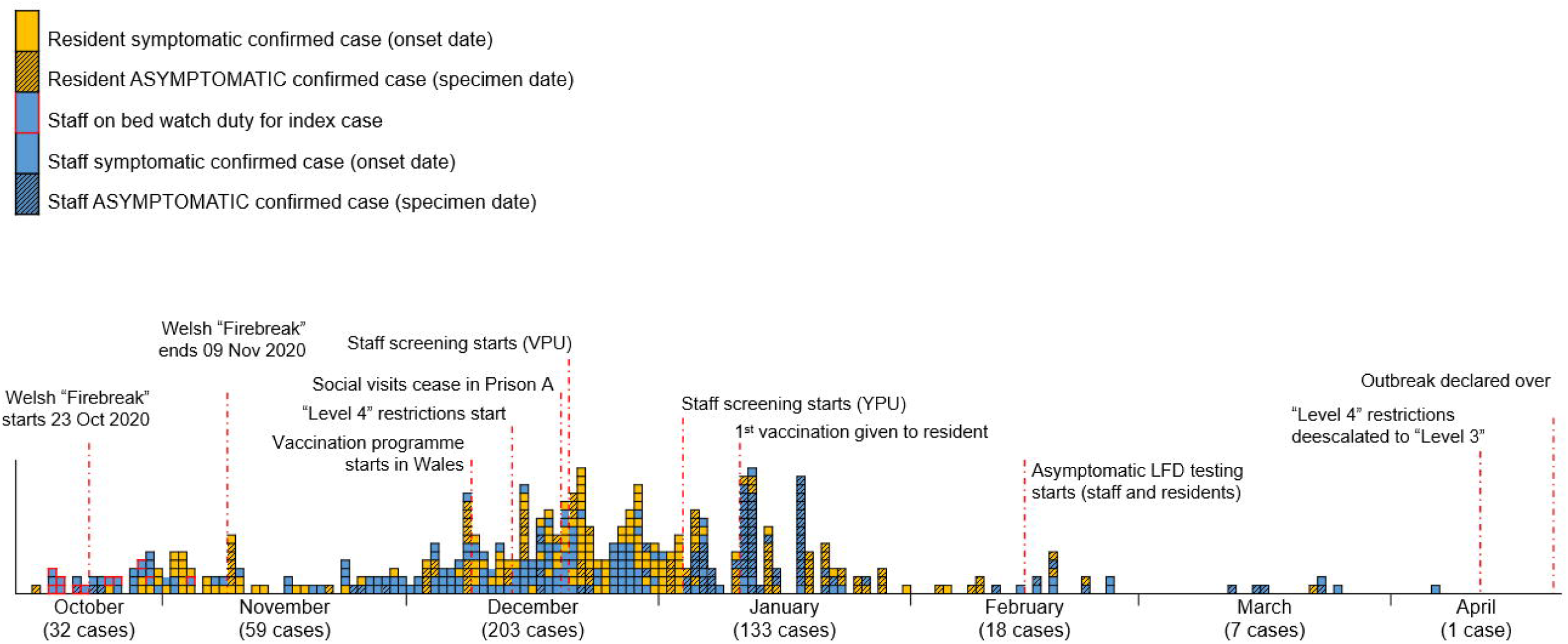

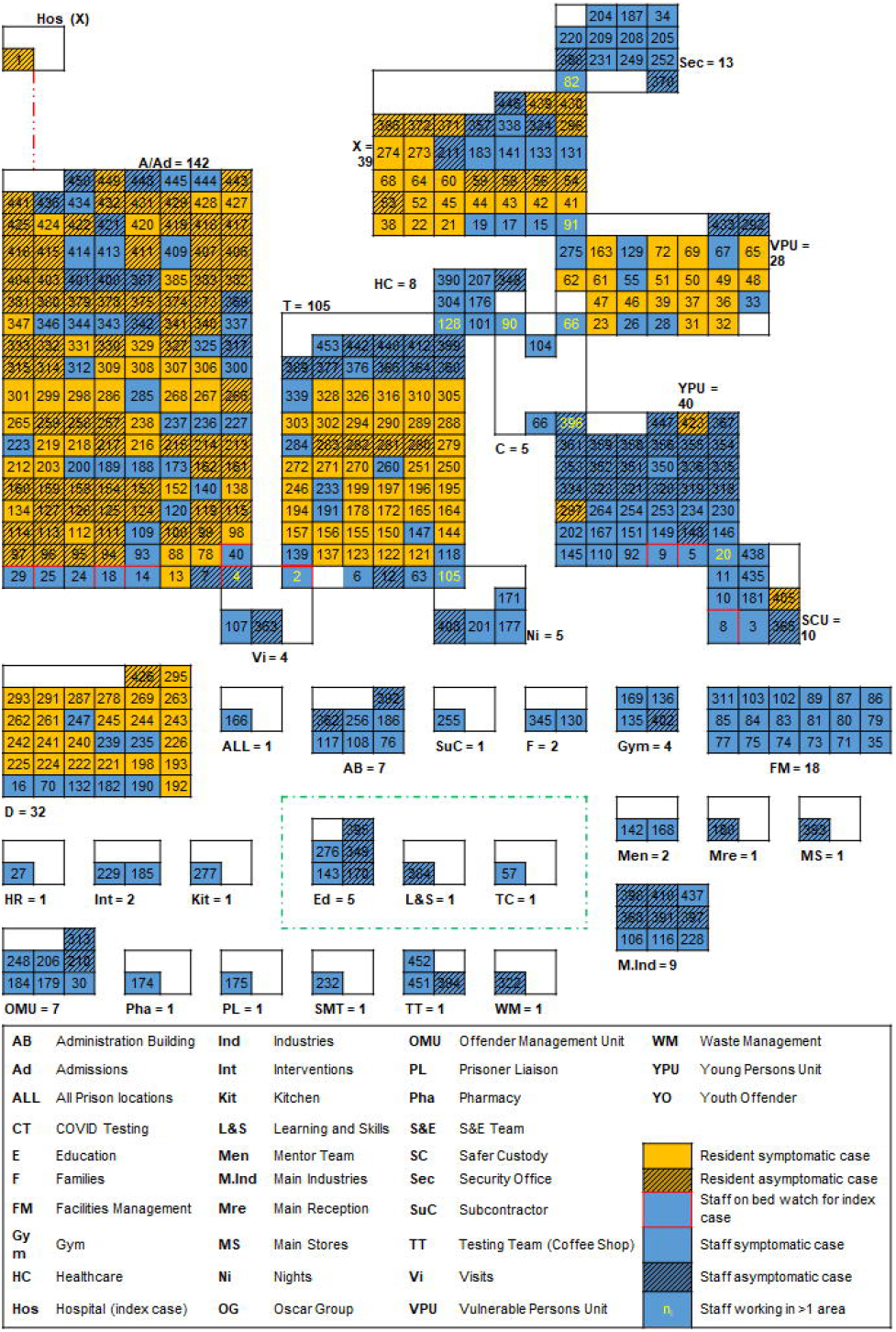

Sequencing of a sample from the index case and one prison staff member on bed watch for the index case revealed that the resident and staff member infections were of different lineages and not closely genetically related.

Subsequent WGS testing showed a third, different phylotype present in the prison; a cluster of seven X-block resident cases employed in the prison’s print shop had identical phylotypes. One staff member’s WGS result was this same phylotype; follow-up interviews revealed brief, informal contact between these cases whilst moving between locations. By the end of October 2020, 26 cases’ test samples had been sequenced (15 residents, 11 staff), identifying in 11 different phylotypes. Phylotypes are based upon the position of a sample on the global SARS-CoV-2 phylogenetic tree, and where two cases possess phylotypes that differ, it is possible to exclude those two cases from being a directly linked transmission. Where two cases possess the same phylotype, when combined with other epidemiological data, it is possible to conclude the cases are part of a transmission group. Taking these WGS results with the other epidemiological data, WGS demonstrated that there had been multiple introductions into the prison as well as subsequent person-to-person transmission.

WGS enabled a delineation of person-to-person spread, and helped demonstrate that this had occurred between residents and staff through identical phylotype results by time, place and person. Person-to-person spread was also found in the same way between staff working in the same location with no contact to residents (Figure 4). Clusters by team, such as in Estates and Facilities Management, were examples of where keeping two metres apart was often difficult when completing essential duties.

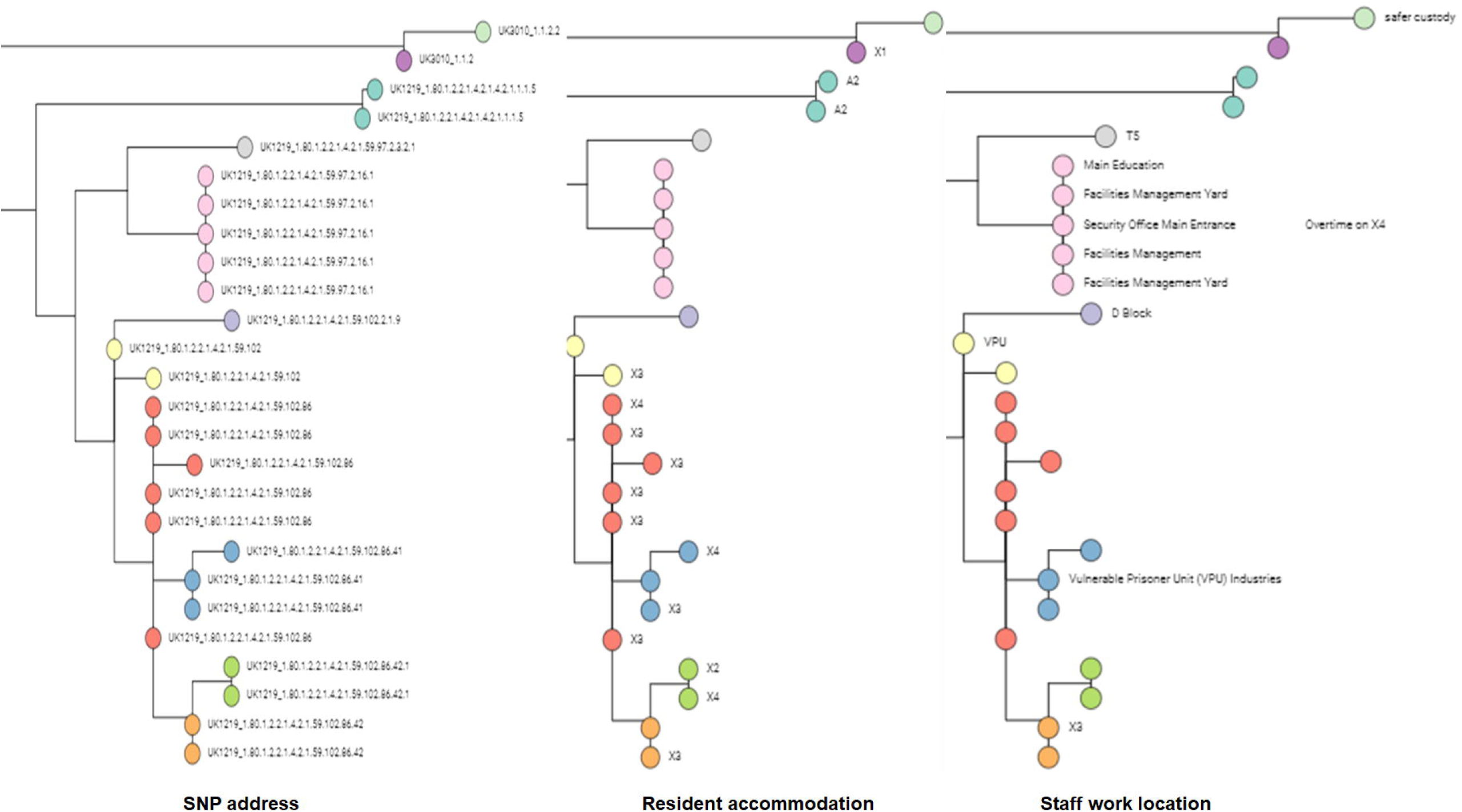

### Outbreak control measures

Certain COVID-19 control measures were in place before this outbreak, mandated across the Welsh Prison estate start of the pandemic:

All areas had enhanced cleaning protocols. Hand-washing stations and 70%-alcohol-gel dispensers were installed in all areas. Safety briefings to all staff and residents reiterated the importance of hand hygiene and social distancing. Facemasks were mandatory for everyone; signage was installed throughout the prison to reinforce these rules. All areas were risk-assessed; rooms stipulated maximum capacity and residents were allowed to move within their blocks in sub-groups to aid social distancing.

Residents were vaccinated by age-cohort by the prison healthcare team; the first vaccination was given in January 2021. Uptake of resident vaccination is affected by prison churn but has been given to 97% of those requesting it (July and November 2021, table 5). Prison staff were offered vaccination in the community by age-cohort, commensurate with the public, which Prison A’s management encouraged.

**Table 5.** Resident COVID-19 vaccination status overview, Prison A (July and November 2021)

|  | July 2021 | November 2021 |
| --- | --- | --- |
| **Total resident population** | **1639** | **1591** |
| Vaccines given | 1555 | 2300 |
| One vaccine dose | 1052 | 1122 |
| Two vaccine doses | 518 | 1090 |
| Refusals | 368 | 464 |
| Waiting to have first vaccine dose | 219 | 21 |

The principal control measures instigated by the OCT are categorised as standard-unilateral measures and targeted-proportional measures. The latter expedited relaxation of other controls imposed based on evidence of reduced infection and transmission:

Staff with COVID-19 symptoms were excluded from work, asked to take a PCR test and remained in self-isolation until the result was known. Symptomatic residents took a PCR test and remained in cell-isolation pending results. Where cells were shared, this contact formed a “bubble”; they followed the same isolation period as their cellmate, dependent on results. New resident-admissions into Prison A, from the courts system or inter-prison transfer, were reverse-cohorted by date to limit transmission in either direction between people living and working in A-block.

Asymptomatic testing (day one and day five PCR tests) was implemented for new resident-admissions from December 2020. Admissions were grouped by admission date until 14 days after their arrival before commencement of the induction-programme and relocation to another permanent residential-block.

To minimise new introductions and person-to-person transmission, staff worked in one area only, unless operational necessity otherwise. Staff overtime was restricted to the same area as normal hours. Staff dining became takeaway during March 2020. Staff were frequently advised not to car share, in-person and by email.

The highest level of restrictions (“level four”) were initiated in mid-December 2020 to control the number of cases at this outbreak’s peak. This included suspension of visits, non-essential work and staff-movement across the prison. Residents were limited to 30-minutes per day to shower and exercise outside cells. Meals and purchases were brought to their cells.

Residents performing essential work (kitchen, cleaning and laundry) were organised into shift-groups so men from the same accommodation unit worked together. Non-essential work was limited or stopped during.

Level four-regime control measures could be lifted when positivity rates were low; to confirm this, asymptomatic testing (PCR) was used on a weekly basis, piloted in staff in the vulnerable prisoners unit (VPU) from December 2020 due to the high attack-rate. Testing was extended to staff in the YPU from January 2021 and offered to all staff and residents later that month. Lateral flow device (LFD) testing began in February 2021, becoming available to all staff and residents by the end of March 2021.

### Impact of the control measures

Effective outbreak management required a collaborative, multi-agency OCT that acted at all policy and operational levels, gathered accurate information and implemented appropriate and timely control measures. Cohorting and testing new-admissions reduced their ability to infect others. Asymptomatic screening identified cases who would otherwise have remained an infection risk and highlighted additional efforts required to end level four regime measures safely in specific areas. The timeline of the outbreak and key dates and control measures is shown in detail in Figure 5.

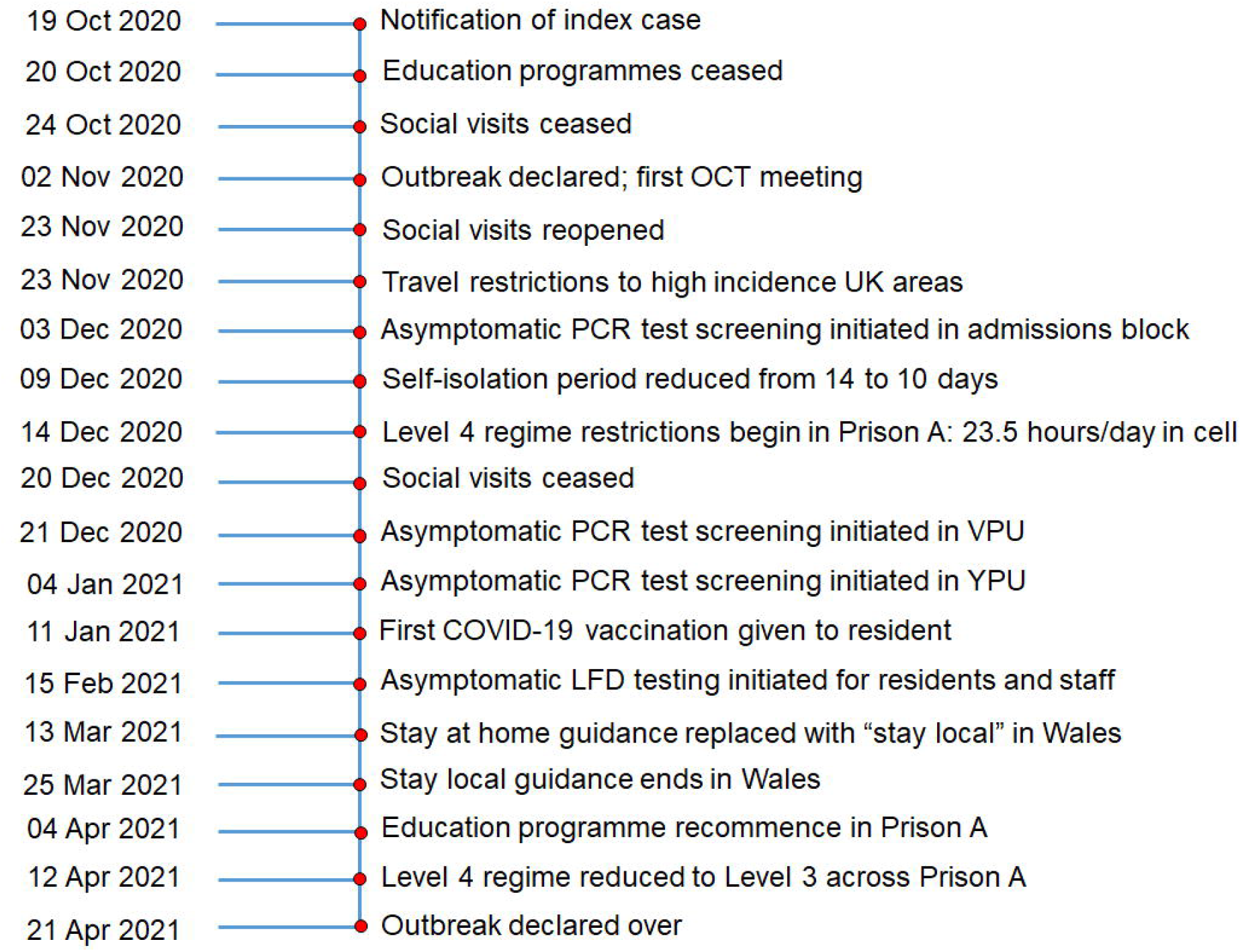

## Discussion

We describe a large, long-running outbreak of COVID-19 affecting 211 prison residents and 242 staff. During the second peak in Wales in December 2020, there were 102 resident cases and 101 staff cases in this outbreak, representing a period-incidence of 60.4 cases per 1000 population (residents) and 118.8 cases per 1000 population (staff) respectively. This was higher for staff but lower for prisoners compared to the England and Wales prison average in the same period, highlighting the impact of community incidence on introductions to the prison. The proportion of cases hospitalised was six-times higher in staff than residents, highlighting potential differences in self-perception of disease severity or inequity of access to healthcare.

There were notable differences in infection patterns; YPU cases were mostly staff (38/40; 95%) and both resident infections were asymptomatic, consistent to community infections whereby young people generally experienced milder or no symptoms of COVID-19. Conversely, D-block cases were mostly residents (24/32; 75%) which may highlight difficulties in social-distancing for those with disabilities. The accommodation with the highest attack rates (SCU=45.5%; YPU=38.5%; D=30.5%) were all self-contained units with fewer residents and staff; thus each new case contributed a higher weighting to attack rates than the larger units.

Where staff had greater exposure to others, attack rates were higher; exemplified by the Mentor and Testing Teams experiencing attack rates of 50%. These roles involved contact with a many of people and, for the Testing Team, an obvious and repeated risk of contact with infectious asymptomatic cases. Because some staff-teams were small and others worked in more than one area, attack rates should be interpreted with caution. What is clear is that the 242 staff cases in a stable population of 850 versus 211 resident cases in a fluctuating population of over 1690 during this outbreak, means the attack rate was far higher in staff than residents (Overall AR = 29% in staff versus 12% in residents). The skew in male cases reflects the resident population and majority of staff being male. The higher proportion of staff cases in this outbreak reached statistical significance. One explanation for this could be the long periods of resident cell-isolation and greatly reduced contact opportunities during socialisation times. Furthermore, staff mixed with others outside work, which might not have been detected by this investigation, despite interviewing.

Controlling COVID-19 outbreaks in prisons requires preventative and reactive measures. This OCT benefitted from dedicated epidemiological support, line listing management and case distribution mapping. The importance placed on epidemiological investigation by the OCT chair improved its ability to determine and expedite control measures through combination of case-location mapping and WGS results. Mapping revealed where further investigation and control measures were required and allowed phased relaxation of restrictions such as cell-isolation. WGS showed this outbreak was seeded by multiple introductions to the prison but also involved person-to-person transmission. In terms of the initial X-block cluster identified by WGS, the same phylotype was widely distributed in communities in Wales at this time in the pandemic and many staff were resident in these areas, giving rise to a plausible introduction-pathway.

Early or temporary release of prisoners was discussed widely early in the pandemic [16], although little evidence suggests this happened at scale anywhere. Cohorting and reduced mixing worked well in this outbreak. However, residents undertake essential services (cooking, laundry, cleaning) resulting in mixing. Prisons have a continuous churn of residents; despite limiting transfers during the peak incidence of COVID-19 in the UK, they were not stopped, resulting in residual risk of infection. Mass screening is effective in identifying cases and limiting spread of disease [17] and was employed here to minimise transmission between groups and identify cases who would have otherwise have remained an infection risk. Scaling-up testing capacity to all staff and residents demonstrated the majority of cases were asymptomatic in the second-half of this outbreak (106 cases in 86 days, 18 symptomatic). If other asymptomatic cases had been identified in this was at the start of this outbreak, the length might have been reduced. It also highlights inequalities in testing practice. This suggests the epidemic curve is not a true reflection of case rates by location, especially in the first-half of this outbreak. The collaborative OCT approach allowed them to discuss, consider, adapt and react within the changing guidance-landscape of the COVID-19 pandemic [18].

Prisons, by design, control activities and movement but provide little personal space. Compared to the wider community, this complicated the control of COVID-19 spreading in a naïve population. As such, stringent social distancing measures were deemed necessary to control COVID-19 in Prison A.

Integration of epidemiological techniques with WGS allowed the OCT to make balanced risk assessments with clarity about direction and intensity of transmission events. As case-rates were reduced in a particular location, relaxation of control measures followed. The social distancing and enhanced hygiene campaigns in place for eight months before this outbreak clarified the impact of the OCT’s control measures.

Vaccination is a key control measure strategy for many infectious diseases. There was much debate about vaccination prioritisation for prison residents and staff given the high propensity for transmission. Modelling has shown that vaccinating everyone living and working in prisons is the most effective strategy for reducing COVID-19 cases, transmission and outbreaks, with an 89% reduction in cases over three years [2]. Combination of high vaccine coverage with other non-pharmaceutical interventions has been shown to reduce cumulative infections by up to 54% in another model [19].

Our methods had several limitations. Different testing pathways for staff and residents limited the number of samples available for WGS, particularly early in this outbreak. Additional, early WGS intelligence might have identified other transmission routes, especially in terms of community introduction via staff and inter-prison introduction via A-block. However, there were huge pressure on WGS services due to the COVID-19 pandemic, and it wasn’t until January 2021 that issues around accessing and sequencing Welsh Pillar 2 samples tested by Lighthouse Laboratories were largely resolved. We only considered activities and movements of staff and residents inside Prison A; it is likely that staff socialised outside Prison A, potentially in contact with asymptomatic cases. Telephone interviews with staff cases investigated plausible vehicles of transmission, such as household contacts, but asymptomatic spread in their communities was not investigated and would have been difficult to measure. Despite agreement of the OCT members, not interviewing resident cases was an information bias and could have revealed additional information relevant to the investigation and control measures. Lack of ascertainment of resident denominators by accommodation area due to prison churn meant comparing staff and resident attack rates was not regularly possible.

We conclude that sufficient epidemiological capacity to investigate outbreaks thoroughly allows OCTs to assess changes and implement appropriate control measures. Traditional epidemiological investigations can be augmented with WGS to determine the phylogeny of infections and inform the epidemiological plausibility of community versus prison transmission. Analysis of epidemiological investigation findings revealed that admissions-block was a persistent source of infections; particular attention should be given to admissions screening. Case-distribution location mapping tracked infection progression, visualising data and allowing easy interpretation to make timely control measure responses. Mapping was equally informative for deciding when to relax control measures safely to improve resident welfare.

Limiting staff movements to one area negatively affected prison operational capacity as more staff became absent due to infection, but helped reduce further transmission. Frequent communication of COVID-19 infection control measures to all staff and residents built an inclusive culture of behaviour applicable to all staff and residents.

COVID-19 is likely to be in circulation for several years. Given the vulnerability of prison residents to infectious diseases, prioritisation of prisoners in COVID-19 vaccination campaigns should be considered.

### Recommendations

1. Sufficient capacity for thorough epidemiological investigations is important for providing timely information for the OCT to determine actions
2. Data visualisation through case-distribution location mapping improves speed of interpretation, aiding risk assessment to determine control measures
3. WGS is a powerful tool in assessing the plausibility of transmission chains and should be introduced as soon as possible in prison outbreak investigations
4. Cohorting, particularly in admissions blocks, is effective in limiting transmission of COVID-19 in prisons
5. Asymptomatic testing is highly effective in identifying cases and can limit transmission events by having them isolate

## Data Availability

The data used in this investigation contain personal identifiable information of a vulnerable population. Anonymised information, including that contained in the supplementary information, required to reproduce these results is available from the corresponding author on reasonable request.

## Acknowledgements

None

## Ethics

Ethical oversight of the project was provided by the PHW Research and Development Division. As this work was carried out as part of the health protection response to a public health emergency in Wales, using routinely collected surveillance data, PHW Research and Development Division advised that NHS research ethics approval was not required. The use of named patient data in the investigation of communicable disease outbreaks and surveillance of notifiable disease is permitted under PHW’s Establishment Order. Data were held and processed under PHW’s information governance arrangements, in compliance with the Data Protection Act, Caldicott Principles and PHW guidance on the release of small numbers. No data identifying protected characteristics of an individual were released outside of the OCT.

## Box 1

Prison A accommodation description

There are approximately 1,700 male residents, consisting of adult and young offenders aged 16 years and over. It has 850 staff, of whom approximately 375 are Prison Custody Officers with the balance made up of support, administration and teaching staff. Prisoners are made up of those on remand, short-term, long-term and life sentences. The residents’ accommodation is as follows:

- **A Block (Admissions):** where the majority of new residents stay, known as “receptions”. There are four units housing around 95 prisoners each
- **B Block:** four separate units housing around 95 prisoners each
- **C Block:** learning needs and disability unit housing approximately 75 self-contained prisoners
- **D Block:** (drug unit) housing around 95 self-contained prisoners
- **E & G Blocks:** The young persons’ unit housing around 60 15-17 year olds, separated from other units with its own gym and education facilities. It has separate staff from other units. Prisoners from this unit do not mix with adult prison units
- **H Block:** The safer custody unit housing around 10-20 people in a self-contained unit
- **T Block:** comprises six separate units housing around 400 prisoners
- **X Block:** separate from main prison with three self-contained wings housing around 350 prisoners. It has its own industries, education and gym facilities. Each unit has its own meal tables, kitchen area and association area
- **\*The vulnerable prisoners Unit:** is comprised of X3 and T6, both separate from each other and other prison units

